# A Novel Diagnostic Test to Screen SARS-CoV-2 Variants Containing E484K and N501Y Mutations

**DOI:** 10.1101/2021.03.26.21253712

**Authors:** Yanan Zhao, Annie Lee, Kaelea Composto, Marcus H. Cunningham, Jose R. Mediavilla, Samantha Fennessey, André Corvelo, Kar Fai Chow, Michael Zody, Liang Chen, Barry N. Kreiswirth, David S. Perlin

## Abstract

Spike protein mutations E484K and N501Y carried by SARS-CoV-2 variants have been associated with concerning changes of the virus, including resistance to neutralizing antibodies and increased transmissibility. While the concerning variants are fast spreading in various geographical areas, identification and monitoring of these variants is lagging far behind, due in large part to the slow speed and insufficient capacity of viral sequencing. In response to the unmet need for a fast and efficient screening tool, we developed a single-tube duplex molecular assay for rapid and simultaneous identification of E484K and N501Y mutations from nasopharyngeal swab (NS) samples within 2.5 h from sample preparation to report. Using this tool, we screened a total of 435 clinical NS samples collected from COVID patients at 8 hospitals within the Hackensack Meridian Health network in New Jersey. While B.1.351 and P.1 variants were absent from the current study, our data revealed a dramatic increase in the frequency of E484K over time, underscoring the need for continuous epidemiological monitoring.

---

The COVID-19 pandemic caused by severe acute respiratory syndrome coronavirus 2 (SARS-CoV-2) has plagued human society causing immeasurable losses in an unprecedented way. In the circumstances, many monoclonal and polyclonal antibodies have been approved for clinical use and several vaccines have been licensed with two mRNA vaccines and one viral vectored vaccine being rolled out for population vaccination in various countries and geographical areas. Opposed to these encouraging progresses and facts, this virus has quickly adapted to various pressures from antiviral therapy and host immunity and evolved independently into several SARS-CoV-2 variants of concern (VOC). These variants, including B.1.1.7 (a.k.a. 501Y.V1), B.1.351(a.k.a. 501Y.V2), and P.1(a.k.a. 501Y.V3) variants, are concerning because they either resist neutralizing antibody and possibly reduce vaccine efficacy or show increased transmissibility, via making some key mutations in the spike protein [1, 2, 3, 4, 5]. Studies have shown that currently observed resistance to neutralizing antibody is largely associated with the E484K mutation [1, 2, 5, 6, 7]. Previously, E484K was only harbored by B.1.351 and P1 variants. The most recent report found that the E484K has been successfully incorporated into some isolates of the B.1.1.7 variant [8]. Moreover, the new variant of interest discovered in New York (B.1.526) also carries the E484K mutation, and alarmingly, fast-spreading over the past two months [9, 10]. Another key spike mutation, N501Y, present in all three VOCs is considered to enhance the binding between spike and the ACE2 receptor in human cells, thus contributing to increased transmissibility and possibly virulence as well [11, 12, 13]. As we continue to understand the impact of these variants on the mode of the ongoing pandemic, efficient and continuous monitoring of these variants is critical to implementation of fast and effective countermeasures to eventually defeat this devastating disease. The closer this genotyping can happen to the testing, the quicker the data can be used and actioned. To this end, we developed a novel molecular diagnostic assay capable of identifying the signature mutations within 2.5 h from sample preparation to report, and used this tool to screen clinical nasopharyngeal swabs (NS) collected from COVID patients at Hackensack Meridian Health (HMH) network hospitals from late December 2020 to February 2021.

This novel genotyping is based on the thermal dynamic difference of molecular beacon (MB) binding with perfectly complementary target or mismatch target. To generate single-stranded target DNA for the MB probe, an asymmetric reverse transcription (RT)-PCR assay was developed to amplify the mutation hotspot region covering both 484 and 501 codons of the S gene (**supplemental material**). Upon completion of thermal cycling, a melting curve analysis is performed to characterize dissociation between the single-stranded DNA product and two differentially labeled MB probes, to enable simultaneous genotyping at both loci. Owing to the probe design, the wildtype (WT) template is expected to generate a higher melting temperature (*Tm*) than that of the mutated genotype at corresponding locus. As shown on **Fig. 1**, E484 WT is featured for a *Tm* at 54.85°C ± 0.19°C, ∼ 5°C higher than the *Tm* of E484K (49.81°C ± 0.07°C). Similarly, the signature *Tm* for N501 WT was 59.97°C ± 0.09°C, higher than 54.78°C ± 0.12°C for N501Y. In a blinded fashion, this test correctly genotyped RNA samples extracted from 6 different reference viral strains, including one WT (SARS-CoV-2 USA WA1/2020), two B.1.1.7 variants (SARS-CoV-2 hCoV-19/USA/CA_CDC_5574/2020 and SARS-CoV-2 hCoV-19/England/204820464/2020), and two B.1.351 variants (SARS-CoV-2 hCoV-19/South Africa/KRISP-EC-K005321/2020 and SARS-CoV-2 hCoV-19/South Africa/KRISP-K005325/2020) purchased from BEI resources, and one E484K variant isolate recently obtained from our network hospital. The analytical sensitivity of the assay was evaluated against 10-fold serial dilutions of RNA prepared from each of the reference viral strain. The assay can reliably identify as low as 200 copies of 484WT, 200 copies of E484K, 20 copies of 501WT, and 200 copies of N501Y per reaction, respectively.

**Figure 1.**
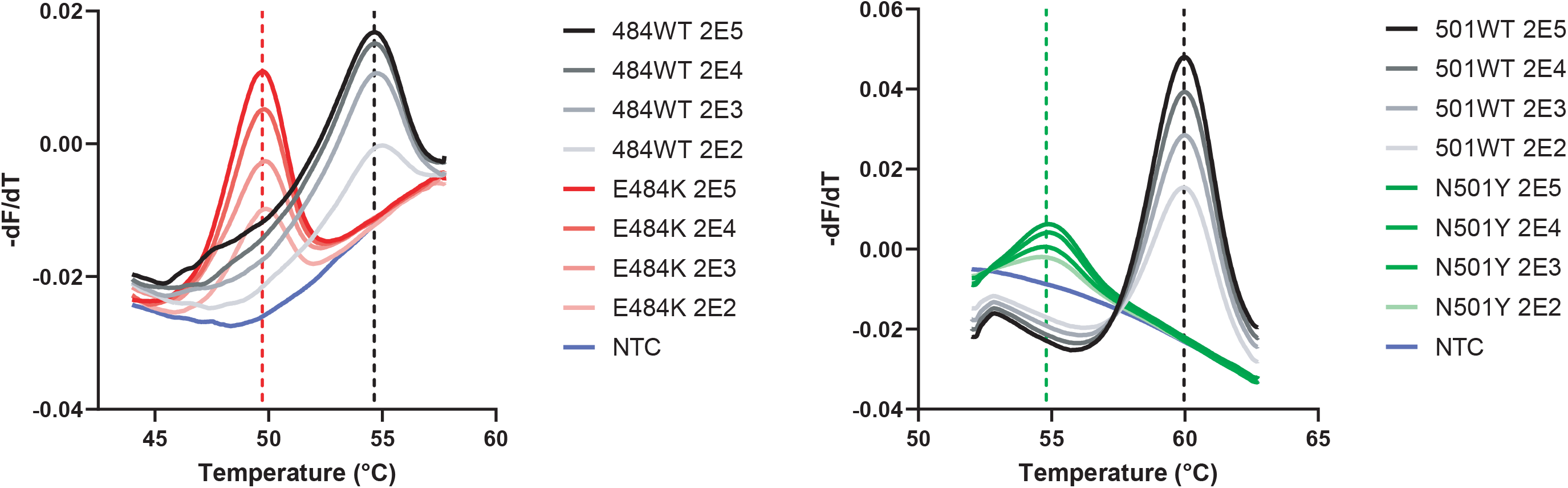
Melting profiles for E484WT and E484K (left panel), and those for N501WT and N501Y (right panel). For each genotype, 10-fold serial RNA dilutions containing 2×10^5^ to 200 genome equivalents/reaction were tested. Dashed lines indicate the *Tm* value of corresponding genotype. NTC is the abbreviation for no template control.

Using this tool, we next accessed to SARS-CoV-2 positive NS specimens through the HMH institutional Biorepository under an HMH-IRB approved protocol Pro2018-1022, to screen the signature mutations. This study also accessed de-identified data for time and location of sample collection through the HMH-IRB approved protocol, Pro2020-0342. A total of 435 samples received in February 2021 from 8 HMH hospitals with a cycle of threshold (Ct) value <37 in SARS-CoV-2 N2 RT-PCR test [14] were subjected to the spike mutation screening.

To speed up the screening procedure, we adopted an extraction-free sample process method by heat inactivating a 50 µl aliquot of swab specimen in the presence of proteinase K at 95°C for 5 min [15], prior to genotyping test. As a result, 365 and 376 samples yielded identifiable signals for 484 and 501 sites, respectively. The proportion of E484K was 8.8% (32/365), and it was 8.0% (30/376) for N501Y. Of note, the presence of E484K and N501Y in the tested samples was completely independent. Co-harboring of both mutations was not observed in our screening. In addition, we discovered a new genotype at the 501-probe binding site from 11 samples (11/376, 2.9%), which thereafter was confirmed to be a N501T (AAT>ACT) mutation in subsequent sequencing. The melting profile of N501T is markedly different from that of WT and N501Y, with a signature *Tm* of 56.41°C ± 0.15°C. A sharp increase was noticed in both E484K and N501Y prevalence in February compared to earlier months. E484K cases account for 12.0% (29/242) of all samples in February. In contrast, it was not detected in samples of December (n=28), and only 3.2% (3/95) of samples of January were E484K positive. The N501Y followed the same trend, with 11.2% (28/251) prevalence in February, and 1.0% (1/98) and 3.7% (1/27) in January and December, respectively. Further analysis by sample collection source found that E484K scattered in various hospitals, whereas N501Y seemed to be highly concentrated in one hospital area (20/107 vs. 10/249, *P*=0.001), indicating a possible small outbreak which warrants further investigation.

We next performed whole genome sequencing (WGS) with a panel of 74 samples representing different genotypes flagged by this screening tool, including 24 E484K, 25 N501Y, 5 N501T, and 20 WT at 484 and 501 loci. Indeed, WGS results are 100% consistent with our genotyping determination. Genomic analysis (**Fig. S**) showed that the majority of the E484K cases (n=19) fell within the B.1.526 lineage, a recent clone emerged from New York, and the rest belong to clade 20C B.1 lineage (n=2), and clade 20B under R.1 (n=2) and B.1.1.309 lineage (n=1), respectively. All N501Y cases except one are members of B.1.1.7 lineage.

Currently, WGS is being used as the main tool for epidemiological monitoring of SARS-CoV-2 variants. However, the relatively long turn-around-time for WGS and the demand on bioinformatic expertise for data analysis makes identification of concerning variants lagged far behind laboratory COVID diagnosis. Clearly, there is an unmet need for faster and simpler screening tool that can be used in a high-throughput fashion to increase the capacity of SARS-CoV-2 variant detection in real-time. Herein, we demonstrate that a novel and easy molecular diagnostic assay can be used as a convenient tool for large scale of SARS-CoV-2 variant screening, thus, to enable highly efficient epidemiological monitoring. Not only the assay is proved to be highly accurate in our clinical screening, notably, it is also sensitive to new mutations within the probe binding site. This has been well exemplified by our discovery of the N501T mutation from the clinical specimens. To our knowledge, this is the first report of using novel screening tool to identify key mutation harboring SARS-CoV-2 variants in NJ hospitals.

B.1.351 and P.1 variants were absent from the current study, however our data revealed a dramatic increase in the frequency of E484K over time, underscoring the need for continuous epidemiological monitoring.

A couple of limitations of this study need to be noted. Firstly, this novel screening assay is slightly less sensitive than the widely used RT-PCR COVID diagnostic assay. Therefore, samples with very low viral load (eg: N2 Ct >35) may not yield identifiable signal in this genotyping test. Efforts towards improving the sensitivity of the test is ongoing. Secondly, the current assay only aims at picking up mutations at 484 and 501 loci. It is our intention to keep expanding and updating this test by adding new mutations associated with important phenotypic change, to better respond to the fast-evolving situation.

## Data Availability

not applicable

## ACKNOWLEDGMENTS

This work was supported by the COVID Emergency Research Fund #61315, Hackensack University Medical center; by funds provided to the CDI by Activision Publishing Inc, Suez North America, and by *NJ Stands Up to COVID*. The funders of this study had no role in the study design, data collection, data analysis, data interpretation, or writing of the report. All authors declare no conflict of interest in this work. Whole genome sequencing was performed at the New York Genome Center (NYGC) as part of the COVID-19 Genomic Research Network (CGRN) with funds generously provided by NYGC donors.

We thank Steven Park, Nathaly Cabrera, and Timothy Mikulski for technical support of viral culture and RNA extraction.

