## Supplemental material for "A Novel Diagnostic Test to Screen SARS-CoV-2 Variants Containing E484K and N501Y Mutations"

**Viral culture and RNA extraction.** Viral culture stocks of SARS-CoV-2 USA WA1/2020 (WT), SARS-CoV-2 hCoV-19/USA/CA_CDC_5574/2020 (B.1.1.7), SARS-CoV-2 hCoV-19/England/204820464/2020 (B.1.1.7), SARS-CoV-2 hCoV-19/South Africa/KRISP-EC-K005321/2020 (B.1.351), and SARS-CoV-2 hCoV-19/South Africa/KRISP-K005325/2020 (B.1.351) were obtained from BEI Resources, NIAID (Manassas, VA). One mutant carrying E4848K but not N501Y was recently isolated and obtained from a HMH network hospital. All strains were propagated on VeroE6/TMPRSS2 cell line (SEKISUI XenoTech, KS) in a Biosafety level (BSL) 3 laboratory. Supernatant of the viral culture was proteinase K treated (200 µg/ml) and heat inactivated at 95°C for 10 min prior to RNA isolation in the BSL-2 laboratory using QIAamp viral RNA mini kit (Qiagen, MD).

**Asymmetric RT-PCR and Melting curve analysis.** One set of primers were designed to amplify a 148-nt region of the SARS-CoV-2 genomic RNA covering E484 and N501 of the spike protein. Two differentially labeled molecular beacons, 484WT-MB (FAM-labeled) and 501WT-MB (Quasar670-labeled), were designed to contain the WT sequences of E484 and N501, respectively. Asymmetric RT-PCR was carried out on the Mic Real Time PCR Cycler (Bio Molecular Systems, software micPCRv2.8.13) in a 20 µl reaction volume using the One Step PrimeScript™ RT-PCR Kit (Perfect Real Time) (Takara). This duplex assay contained 10 µl of one step RT-PCR Buffer III, 0.4 µl of PrimeScript RT enzyme Mix II, 0.4 µl of TaKaRa Ex Taq HS (5U/µl), 40 nM of S484F, 1 µM of S501R (10 µM), 100 µM of both 484WT-MB and 501WT-MB, and 5 µl of RNA or heat-inactivated template. Thermal cycling profile was 42°C for 5 min for reverse transcription, followed by 95°C for 10 sec then 50 cycles of 95°C for 5 sec and 60°C for 20 sec. Immediately after amplification, melting curve analysis was initiated as a minute incubation at 95°C, after which it was melted from 47.5°C to 58.5°C with a ramp rate of 0.1°C/s for the 484WT-MB and melted from 53°C to 63°C with a ramp rate of 0.1°C/s for the 501WT probe.

**Viral genome sequencing and analysis**. Viral RNA from swabs was extracted using QIAcube Connect (Qiagen), following the manufacturer's instructions. SARS-CoV-2 targeted assay libraries were prepared using the QIAseq SARS-CoV-2 Primer Panel and cDNA Synthesis for Illumina kits (Illumina). Adapter sequences and low quality (Q < 20) bases were trimmed from the raw reads, using Cutadapt v2.101 (https://github.com/marcelm/cutadapt/). Processed reads were then mapped to the SARS-CoV-2 genome reference using BWA-MEM v0.7.172 [1] and genome sequences were determined by Samtools v1.11 and bcftools v1.11 [2]. The genome clades and lineages were determined by Nextclade server (https://clades.nextstrain.org/) and Pangolin v2.3.0 (https://github.com/cov-lineages/pangolin), respectively. Genomes were aligned using nextalign v0.2.0 (https://github.com/nextstrain/nextclade/releases), and a maximum likelihood phylogenetic tree was constructed using IQ-TREE v2.1.2 [3] with automatic model selection and 1000-bootstrap replicates. The tree was annotated using iTOL v6.0 [4].

**Clinical screening.** Clinical samples included in this study were de-identified nasopharyngeal swabs obtained from 8 different sites within HMH network and banked in the biorepository of our center. All samples were collected in standard viral transport media and stored at -80°C upon receipt. An extraction-free method [5] was used to process samples prior to genotyping analysis. Briefly, a 50 µl of aliquot was taken from each swab and mixed with 6.5 µl of proteinase K (20 mg/ml, Roche), followed by heating up the mixture at 95°C for 5 min. Then 5 µl of the processed sample was used directly as template for genotyping assay. Information of sample source and collection timeline was provided by HMH bio-R working group.

**Statistical analysis.** *Tm* values for E484 and N501 genotype were determined by melting curve analysis using the Mic Real-Time PCR software (micPCRv2.8.13). The melting curve and epidemiological distribution of variants were plotted and analyzed in GraphPad Prism version

9.0.0. We used $\chi^2$ test or Fisher's exact test to compare the distribution between different locations. A *P* value less than 0.05 was considered statistically significant.

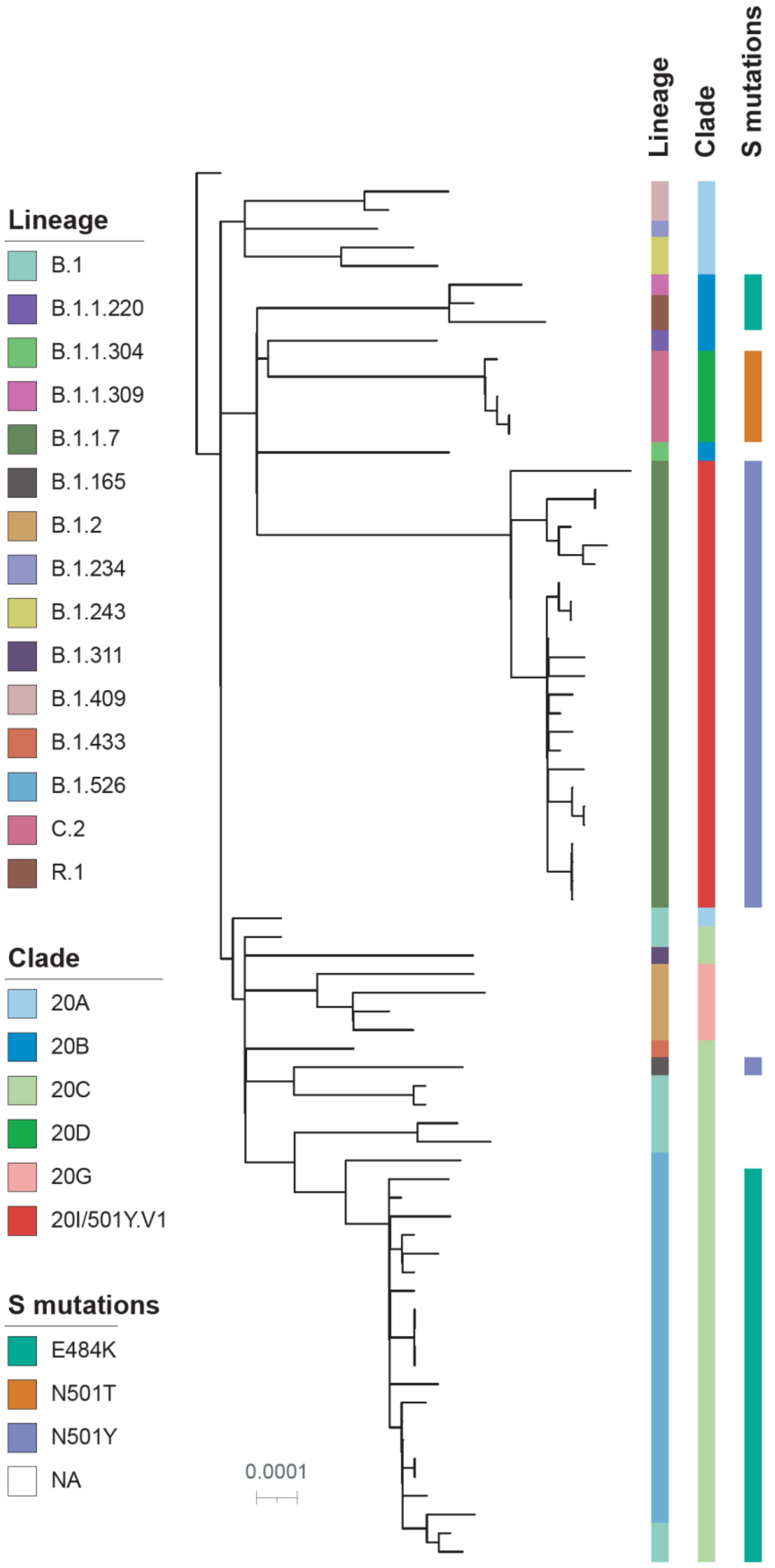
Lineage
Clade
S mutations
Lineage
B.1
B.1.1.220
B.1.1.304
B.1.1.309
B.1.1.7
B.1.165
B.1.2
B.1.234
B.1.243
B.1.311
B.1.409
B.1.433
B.1.526
C.2
R.1
Clade
20A
20B
20C
20D
20G
20I/501Y.V1
S mutations
E484K
N501T
N501Y
NA
0.0001

**Figure S1.** A maximum-likelihood phylogenetic tree of 74 SARS-CoV-2 genomes from HMH network. The tree is rooted with the Wuhan/Hu-1 SARS-CoV-2 reference (NC_045512.2) sequence and annotated using iTOL (www.itol.embl.de). The scale bar represents 0.0001 nucleotide substitutions per site. The SARS-CoV-2 Pangolin lineage, NextStrain clade, S protein 484 and 501 mutations were illustrated by different color bars on the right.
